# FHIR-GPT Enhances Health Interoperability with Large Language Models

**DOI:** 10.1101/2023.10.17.23297028

**Authors:** Yikuan Li, Hanyin Wang, Halid Z. Yerebakan, Yoshihisa Shinagawa, Yuan Luo

## Abstract

Advancing health interoperability can significantly benefit health research, including phenotyping, clinical trial support, and public health surveillance. Federal agencies, including ONC, CDC, and CMS, have been collectively collaborating to promote interoperability by adopting Fast Healthcare Interoperability Resources (FHIR). However, the heterogeneous structures and formats of health data present challenges when transforming Electronic Health Record (EHR) data into FHIR resources. This challenge becomes more significant when critical health information is embedded in unstructured data rather than well-organized structured formats. Previous studies relied on multiple separate rule-based or deep learning-based NLP tools to complete the FHIR resource transformation, which demands substantial development costs, extensive training data, and meticulous integration of multiple individual NLP tools. In this study, we assessed the ability of large language models (LLMs) to transform clinical narratives into HL7 FHIR resources. We developed FHIR-GPT specifically for the transformation of clinical texts into FHIR medication statement resources. In our experiments using 3,671 snippets of clinical texts, FHIR-GPT demonstrated an exceptional exact match rate of over 90%, surpassing the performance of existing methods. FHIR-GPT improved the exact match rates of existing NLP pipelines by 3% for routes, 12% for dose quantities, 35% for reasons, 42% for forms, and over 50% for timing schedules. Our findings provide the foundations for leveraging LLMs to enhance health data interoperability. Future studies will aim to build upon these successes by extending the generation to additional FHIR resources.

## Introduction

Interoperability enhances the ability of healthcare providers to deliver safe, effective, and patient-focused care. It also offers novel avenues for individuals and caregivers to access electronic health data for care coordination and management ^1^. The promotion of interoperability has become an integral aspect of various health initiatives, spanning from ensuring health equity to responding to public health emergencies ^2^. Federal agencies, including the Office of the National Coordinator of Health IT (ONC) ^1^, the Centers for Disease Control and Prevention (CDC) ^3^, and the Centers for Medicare & Medicaid Services (CMS) ^4^, collectively collaborate to promote interoperability through the adoption of FHIR, which is an interoperability standard developed by the Health Level 7 (HL7®) standards development organization.^5^ FHIR is specifically designed to facilitate the swift and efficient exchange of health data. FHIR has seen growing adoption in the modeling and integration of both structured and unstructured data for various health research purposes. Its applications range from developing computational phenotyping ^6-8^ to supporting clinical trials ^9-12^, building surveillance systems ^13,14^, and much more.

Transforming health data into the FHIR format presents a major challenge, as various health organizations have their unique infrastructure, standards, and formats for generating, storing, and organizing health data ^15^. This challenge becomes more significant when critical health information is embedded in unstructured data other than well-organized structured formats. There are existing efforts for promoting the transformation of unstructured data into FHIR resources, offered by both academic and commercial sectors. In academic research, Hong et al.^16^ integrated clinical NLP tools, including cTAKES ^17^, MedXN ^18^, and MedTime ^18^, to extract clinical entities from corresponding document sections and standardize them into FHIR resources. Wang et al. developed Opioid2FHIR ^19^, a system that employs multiple deep learning-based natural language processing (NLP) techniques for opioid information extraction and normalization. In the commercial domain, Google Cloud has released the Healthcare Natural Language API ^20^, capable of converting medical text input into FHIR resources. Azure Health Data ^21^ is proficient at converting semi-structured data into FHIR resources but does not handle free-text unstructured input. All the above FHIR transformation tools necessitate multiple NLP tools in sequence. Creating a pipeline that integrates multiple NLP tools requires substantial computational resources, annotated data, and human efforts. Furthermore, as the transformation progresses along the pipeline, the errors from each NLP tool compound and decrease overall accuracy.

Therefore, we proposed leveraging pre-trained large language models (LLMs) and meticulous prompt engineering to facilitate the transformation of free-text input into FHIR resources. We manually annotated a dataset of free-text to FHIR *MedicationStatement* resource transformation pairs. We compared the transformation accuracy between FHIR-GPT and existing NLP pipelines using the annotated dataset.

## Methods

In this section, we delve into the technical details employed in data annotation, LLMs usage, and the evaluation process. **Error! Reference source not found**. is an illustrative visual representation of the workflow.

To the best of our knowledge, there is no publicly available dataset with corresponding text and FHIR resource pairs. We therefore annotated a dataset containing pairs of free-text input and corresponding FHIR *MedicationStatement* resource output. The FHIR resource of *MedicationStatement* is a record of *a medication being taken by a patient or that a medication has been given to a patient, where the record is the result of a report from the patient or another clinician, or derived from supporting information*.^22^ This transformation holds particular significance because many medication-related details, such as the reasons for administration and dosage instructions, often remain absent in structured data. Clinical notes within the EHR system frequently serve as the primary source for this medication-related information. For detailed examples of the elements in *MedicationStatement*, please refer to **Error! Reference source not found**..

The clinical text input was obtained from the 2018 n2c2 medication extraction challenge.^23^ We extracted the text snippets, each containing mentions of one medication and all its associated entities, from the discharge summaries. These extracted snippets, each tied to a specific medication, serve as input for both annotations and transformations. Our human annotation consisted of three key steps. We started by standardizing the entities from free text into clinical terminology coding systems. To achieve this, we leveraged the word spans of entities provided in the n2c2 dataset and manually looked up the HL7 CodeSets, SNOMED CT Browser ^24^, and the RxNav ^25^ for standardization. We then assembled the identifiers, codes, texts, elements, and structures into a complete *MedicationStatement* resource in JSON format as per FHIR v6.0.0: R6 implementation guide ^22^. Finally, the human-converted *MedicationStatement* resources underwent validation using the official FHIR validator^26^ to ensure compliance with FHIR standards, including structure, datatypes, cardinalities, code sets, display names, etc.

We experimented with the following 3 Large Language Models (LLMs), OpenAI GPT-4 ^27^, Llama-2-70B ^28^, and Falcon-180B ^29^. We accessed the GPT-4 (model: *gpt-4-32k as of 2023-05-15*) through the Azure OpenAI API service. We made multiple asynchronous API calls to enhance efficiency. For Llama-2-70B and Falcon-180B, we deployed them on our HIPAA-compliant firewalled local servers with multiple GPUs. GPTQ ^30^ was used to accelerate the inference time for Llama-2-70B and Falcon-180B.

We required these LLMs to transform the free-text entries into *MedicationStatements* conforming to the FHIR standard, employing the few-shot prompt settings that include 4-5 examples of transformations in the prompts. Each clinical text snippet was individually input into the LLMs to generate their *MedicationStatement* resource. We leveraged five separate prompts to instruct the LLM to transform the free-text input into the elements of a *MedicationStatement* resource, including medication details (drug name, strength, and form), route, timing, dosage, and reason. All few-shot prompts followed a template consisting of task instructions, expected output FHIR templates in JSON format, 4-5 examples of transformations, a comprehensive list of codes from which the model could make selections, and the input text to be transformed. As there was no fine-tuning or domain-specific adaptation in our experiments, we initially had the LLMs generate the FHIR resource for a small subset of the dataset (N=100). Then, we manually reviewed the discrepancies between the LLM-generated FHIR output and our human annotations. Common mistakes were identified and used to refine the prompts. There were slight differences in the prompts for each LLM, as different LLMs may be sensitive to different prompts. We did not instruct the LLMs to look up the SNOMED codes for the ‘medication’ and ‘reason’ elements, as there are thousands of SNOMED CT Medication and Finding codes, exceeding the token limits of LLMs. Instead, our instructions were for them to identify the contexts mentioned in the input text and convert them into the appropriate JSON format. For other elements, such as routes and forms, we instructed LLMs to directly look up from the code set (numbering in hundreds). Example prompts can be found in the supplemental material. We will also share our annotated dataset on physionet.org upon acceptance.

We compared LLMs with two existing NLP pipelines: NLP2FHIR ^16^ and Google HNL API ^20^. NLP2FHIR was built based on a previous version of the FHIR implementation guide R5; the Google HNL API primarily standardized concepts to UMLS CUIs, while the latest guide R6 and SNOMED was used in our annotations and LLMs’ transformations. We therefore made necessary adaptations and conversions to ensure a fair comparison. We deployed the NLP2FHIR pipeline on our firewalled local servers and accessed the Google HNL API through the Google Cloud Healthcare API.

When evaluating the FHIR resources generated by the LLMs, we first conducted a format validation check to ensure that the output was in a valid JSON format. On passing the validation, we evaluated the generated resources with exact match rate. This strict criterion required that the resources generated by the LLMs exactly matched the human annotations in all aspects, including structures, codes, and cardinality.

## Results

The annotation results are presented in **Error! Reference source not found**.. In summary, we annotated a total of 3,671 pairs of free-text input and FHIR *MedicationStatement* resource output. The free-text input was derived from discharge summaries for 280 admissions. The annotated resources encompass 625 distinct medications in 26 different forms and are associated with 354 different reasons, as well as 16 administration routes. These elements display varying levels of availability, ranging from approximately 30% for reasons to 65% for timing schedules. The annotated resources in the JSON structure have an average number of objects of 58.2 (standard deviation = 16.2) and an average depth of 6.7 (standard deviation = 0.5).

The transformation results are presented in **Error! Reference source not found**.. In summary, transformation using FHIR-GPT, achieved an exceptional exact match rate of over 0.90 for all elements, outperforming both baseline models and all other LLMs. Specifically, when compared to existing NLP pipelines, FHIR-GPT improved the exact match rate by 3% for routes, 12% for dose quantities, 35% for reasons, 42% for forms, and over 50% for timing schedules. Among all LLMs, we observed a trend of increasing accuracy as the parameter size increased. FHIR-GPT, with approximately 1.7 trillion parameters, surpassed the 180 billion parameter Falcon model and further improved upon the 70 billion parameter Llama-2 model.

## Discussion

The reproducibility of using LLMs for FHIR transformation was examined through two experiments conducted six months apart, in September 2023 and March 2024. No updates were applied to the weights of Falcon and Llama during this period. In our March 2024 experiment of FHIR-GPT, we employed the latest *gpt-4-turbo* model, an upgrade from the *gpt-4-32k* model utilized in the previous September 2023 run. The *gpt-4-turbo* model boasts an expanded context window, stretching from 32k to 128k tokens, and incorporates additional training data spanning from September 2021 to April 2023. The results of reproducibility can be found in **Error! Reference source not found**.. Although there are slight fluctuations observed across various models and elements, none exceed a decrease of 2 percent. This indicates a relative stability in using LLMs for FHIR transformation, even with the update to the foundation model.

We conducted an error analysis to investigate instances where FHIR-GPT and human annotation diverge, with a particular focus on drug routes as an example. The 204 disagreements in transforming drug routes were categorized into five types of errors: false negatives, false positives, mismatched errors, syntax errors, and content filter rejections. A comprehensive breakdown, along with examples and distribution of these errors, is provided in **Error! Reference source not found**.. False negative errors primarily result from FHIR-GPT’s insensitivity to certain medical abbreviations, such as ‘gtt’, or its failure to associate medical terms like ‘lumen’ with the intravenous route. Conversely, false positive errors occur when FHIR-GPT inaccurately introduces nonexistent information or identifies annotation errors, such as the oversight of ‘IVIG’, which was unrecognized in the i2b2 expert annotation and therefore omitted from our human annotation. The mismatched-error category presents a combination of false negative and false positive errors. We posit that such errors could be mitigated through the incorporation of more domain-specific knowledge or examples in the prompt, or through the injection of knowledge bases with retrieval-augmented generations. Additionally, rare instances occur where output cannot be parsed as JSON. These parsing issues can be easily rectified with a simple format adjustment, replacing all double quotes with single quotes or using more advanced JSON parsing tools. Additionally, the Azure platform occasionally rejects requests with content filters to avoid harmful content in the prompts, though such filters can be opted out of if necessary.

In this study, we delve into three potential pathways for transforming free-text clinical input into FHIR resources. While human annotation is the gold standard for transformation, its dependence on extensive human efforts poses scalability limitations. Existing NLP pipelines can automate these transformation processes but demand substantial training data and resources, with challenges in generalizability and transferability. On the other hand, a new pipeline must undergo training or fine-tuning for even minimal changes in the code set or expansion to new resources. In addition, the multi-step transformation process incurs considerable maintenance costs, demanding meticulous tracing for effective error debugging across all steps. FHIR-GPT, harnessing the power of pre-trained LLMs, eliminates the need for high-cost training and depends on only minimal human annotation for the few-shot examples in the prompts. FHIR-GPT also achieves a superior level of accuracy compared to the approach employing NLP pipelines. Moreover, by altering the template and the corresponding code set in the prompt, FHIR-GPT holds the potential to generalize to other resources without the requirement for resource-specific training or fine-tuning. We believe that leveraging FHIR-GPT has the potential to greatly enhance interoperability, given its ease of implementation, high accuracy, and broad generalizability.

We acknowledge the following limitations in our study. irstly, while FHIR-GPT showcases superior performance compared to Llama and Falcon, its significant computational resource demands present a challenge. Moreover, its commercial nature gives rise to ethical considerations, impeding smooth integration into local EHR systems. We aim to investigate alternative lightweight and open-source foundation models of GPT-4 to overcome these obstacles while upholding comparable performance. Secondly, our evaluation of FHIR-GPT was confined to medication-related FHIR resources, potentially limiting the broader applicability of our findings. To address this, our future efforts aim to expand the transformation for a wider array of FHIR resources. Thirdly, our current approach primarily involves prompt engineering through trial-and-error with existing LLMs. There is no enhancement of the architecture of the foundational LLMs. To improve accuracy and reasoning in the transformation process, we aim to adopt advanced techniques like chain-of-thoughts^31^, retrieval-augmented generation, or domain-specific finetuning in future endeavors. Fourthly, we recognize that LLMs can assist in other FHIR-related transformations, such as FHIR version upgrades and tabular-to-FHIR transformations. While we perceive these tasks as potentially less complex than our text-to-FHIR transformations, we encourage fellow researchers to explore the efficacy of LLMs in these alternative pathways to enhance interoperability.

## Conclusion

In conclusion, this study lays the groundwork for harnessing LLMs to significantly improve health data interoperability through the transformation of free-text input into the FHIR resources. The FHIR-GPT model not only streamlines the process but also enhances transformation accuracy when compared to existing NLP pipelines. Building upon these promising results, our future investigations will expand to encompass additional FHIR resources, aiming to advance the practical applications of LLMs in enhancing health data interoperability.

## Data Availability

https://mimic.mit.edu/docs/iii/
https://portal.dbmi.hms.harvard.edu/projects/n2c2-nlp/

---

**Table 1.**
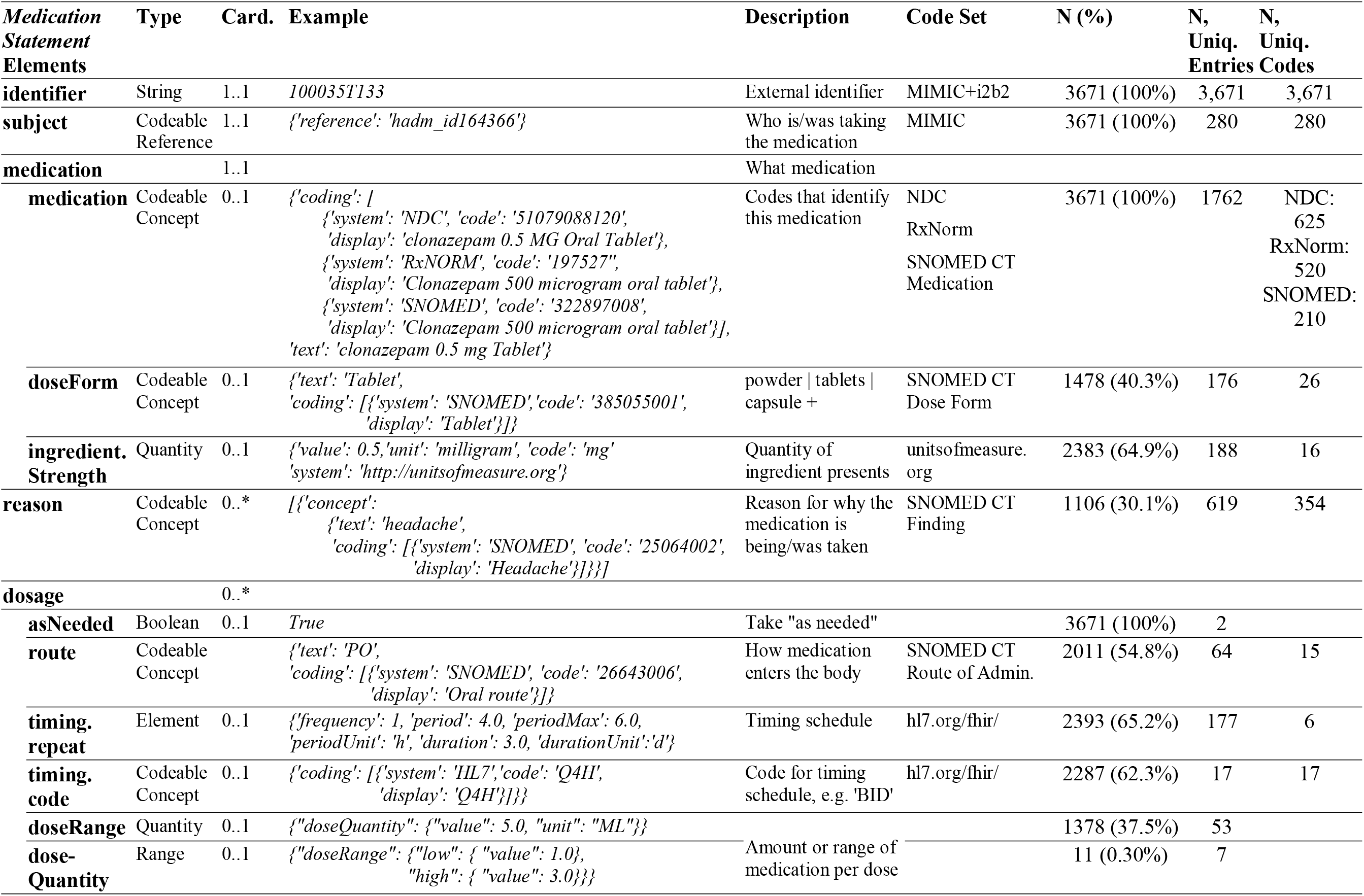
Descriptions, examples, and statistics of human annotation for the FHIR *medicationstatement* resource.

**Table 2.** Comparison of LLMs and existing NLP pipelines for transforming free-text input into FHIR *MedicationStatement* resources. Performance is evaluated using the exact match rate, which requires that the resources generated by the models precisely match human annotations in all aspects, including structure, codes, and cardinality. Due to version and implementation differences, the existing NLP pipelines cannot generate all the elements included in our annotations. The best-performing model for each element is indicated in bold, while the second-place model is underlined.

| Elements of *medicationstatement* | Large Language Models |  |  | Existing NLP Pipelines |  |
| --- | --- | --- | --- | --- | --- |
|  | FHIR-GPT<sup>27</sup> | Falcon-180B<sup>28</sup> | Llama-2-70B<sup>29</sup> | NLP2FHIR<sup>16</sup> | Google HNL API<sup>20</sup> |
| **medication** |  |  |  |  |  |
| **medication** | **0.968** | 0.899 | 0.859 | 0.862 | <u>0.963</u> |
| **doseForm** | **0.976** | <u>0.790</u> | 0.633 | 0.556 | - |
| **ingredient.Strength** | **0.980** | <u>0.921</u> | 0.792 | - | - |
| **reason** | **0.902** | 0.593 | 0.169 | <u>0.645</u> | - |
| **dosage** |  |  |  |  |  |
| **route** | **0.902** | 0.457 | 0.516 | - | <u>0.871</u> |
| **timing.repeat** | **0.947** | 0.268 | 0.221 | 0.403 | - |
| **timing.code** | **0.952** | <u>0.818</u> | 0.600 | 0.424 | - |
| **doseQuantity/Range** | **0.973** | <u>0.864</u> | 0.823 | 0.724 | 0.854 |

**Table 3.** Reproducibility in using LLMs for transforming FHIR resources. Two separate experiments were conducted six months apart using the same prompts, with identical model weights used for Falcon and Llama models. FHIR-GPT used the gpt-4-32k model in the September 2023 experiment, which was upgraded to the gpt-4-turbo (128k) model in March 2024.

| Elements of *medicationstatement* | Large Language Models |  |  |  |  |  |
| --- | --- | --- | --- | --- | --- | --- |
|  | FHIR-GPT<sup>27</sup> |  | Falcon-180B<sup>28</sup> |  | Llama-2-70B<sup>29</sup> |  |
|  | Sept. 2023 | Mar. 2024 | Sept. 2023 | Mar. 2024 | Sept. 2023 | Mar. 2024 |
| **medication** |  |  |  |  |  |  |
| **medication** | 0.968 | 0.970 | 0.899 | 0.897 | 0.859 | 0.849 |
| **doseForm** | 0.976 | 0.974 | 0.790 | 0.785 | 0.633 | 0.634 |
| **ingredient.Strength** | 0.980 | 0.981 | 0.921 | 0.912 | 0.792 | 0.792 |
| **reason** | 0.902 | 0.908 | 0.593 | 0.617 | 0.169 | 0.172 |
| **dosage** |  |  |  |  |  |  |
| **route** | 0.902 | 0.944 | 0.457 | 0.471 | 0.516 | 0.518 |
| **timing.repeat** | 0.947 | 0.938 | 0.268 | 0.264 | 0.221 | 0.218 |
| **timing.code** | 0.952 | 0.946 | 0.818 | 0.810 | 0.600 | 0.580 |
| **doseQuantity/Range** | 0.973 | 0.972 | 0.864 | 0.864 | 0.823 | 0.814 |

**Table 4.**
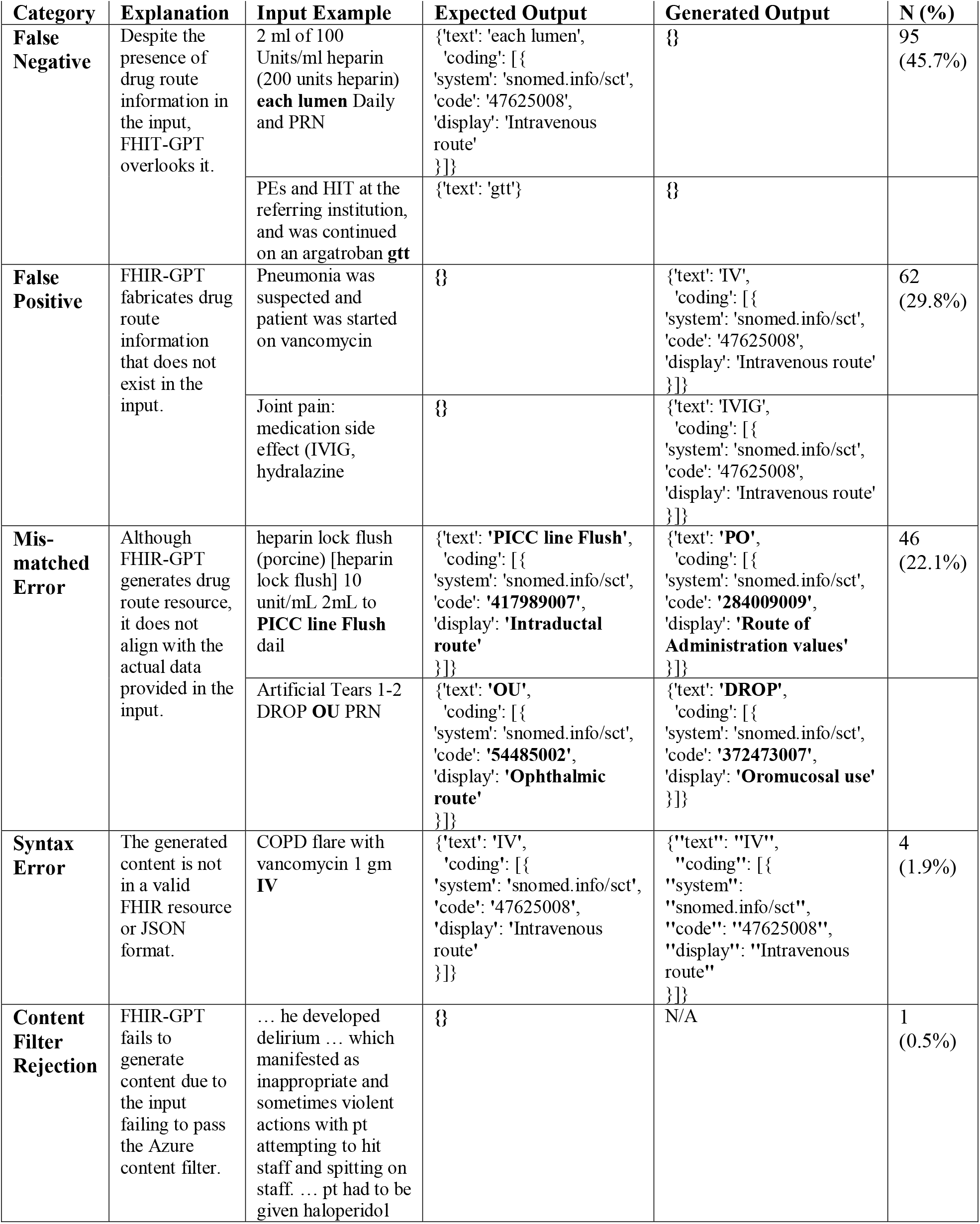
Discrepancies between FHIR-GPT generated resources and human annotations. 204 disagreements in transforming drug routes were categorized into five types of errors.

**Figure 1.**
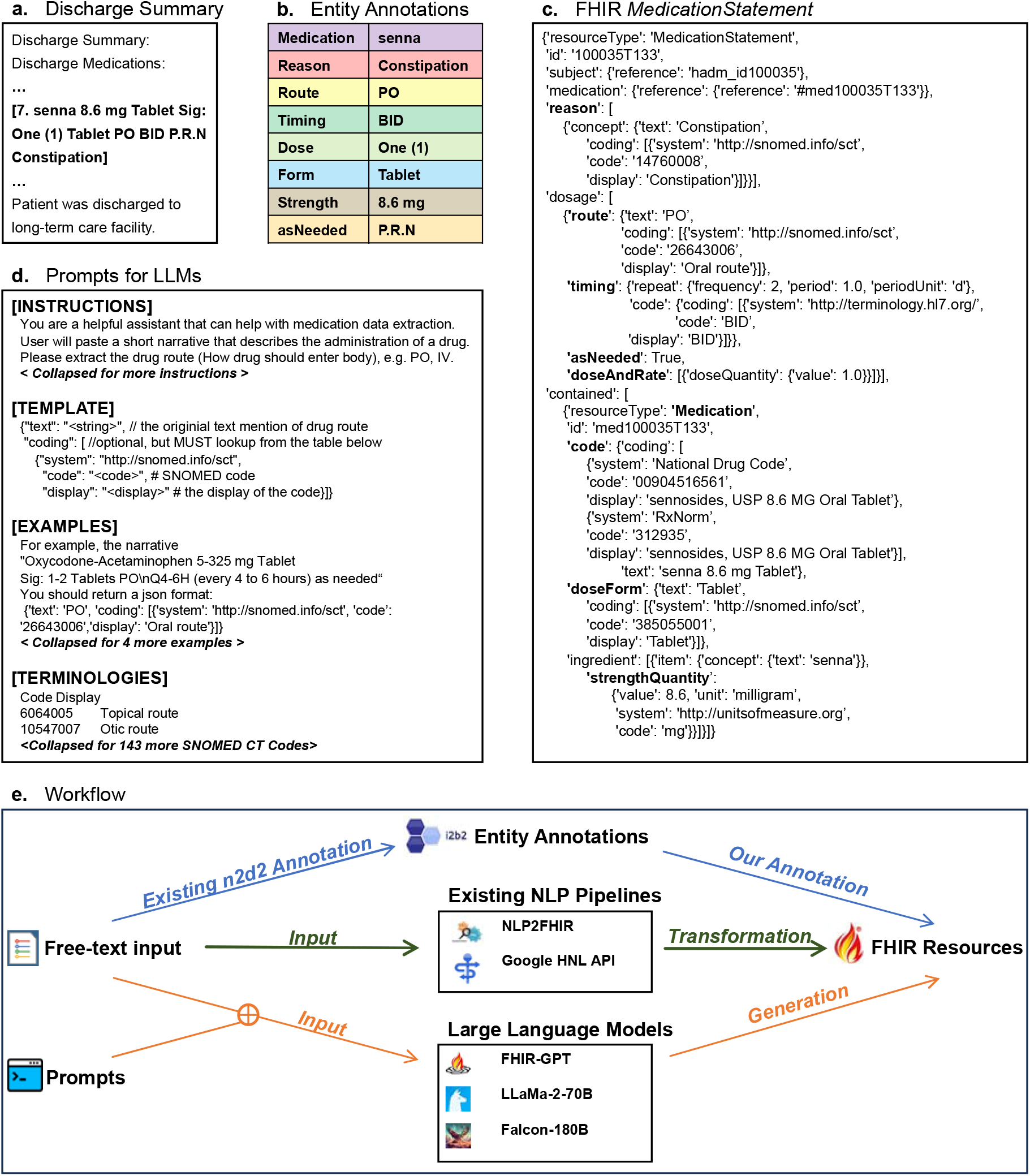
Overview of the transformation from free-text to FHIR resource. **a**. Example of a snippet from the discharge summary, which is the free-text input for FHIR resource generation. **b**. The n2c2 expert annotation of medication-related entities in the discharge summary. **c**. Example of the transformed FHIR *MedicationStatement* resource based on our annotations, serving as the ground truth transformation. The same color shading from panel b is used. **d**. Example prompt used to instruct LLMs in generating FHIR resources. **e**. The workflow details how we annotated the dataset and compared the performance of LLMs with existing NLP pipelines in transforming free-text inputs into associated FHIR resources.]

